# Prediction of the time evolution of the Covid-19 Pandemic in Italy by a Gauss Error Function and Monte Carlo simulations

**DOI:** 10.1101/2020.03.27.20045104

**Authors:** Ignazio Ciufolini, Antonio Paolozzi

## Abstract

In this paper are presented predictions on the evolution in time of the number of positive cases in Italy of the Covid-19 pandemic based on official data and on the use of a function of the type of a Gauss Error Function as a Cumulative Distribution Function (CDF). We have analyzed the available data for China and Italy. The evolution in time of the number of cumulative diagnosed positive cases of Covid-19 in China very well approximates a distribution of the type of the Error Function, that is, the integral of a normal, Gaussian distribution. We have then used such a function to study the potential evolution in time of the number of positive cases in Italy by performing a number of fits of the official data so far available. We then found a statistical prediction for the day in which the peak of the number of daily positive cases in Italy occurs, corresponding to the flex of the fit, i.e., to the change in sign of its second derivative (that is the change from acceleration to deceleration) as well as of the day in which a substantial attenuation of such number of daily cases is reached. We have then performed 150 Monte Carlo simulations in the attempt to have a more robust prediction of the day of the above-mentioned peak and of the day of the substantial decrease of the number of daily positive cases. Although, official data have been used, these predictions are obtained with a heuristic approach, since those predictions are based on statistical approach and do not take into account either a number of relevant issues (such as medical, social distancing, virologic, epidemiological, etc.) or models of contamination diffusion.

## 1. Introduction

By considering the cumulative diagnosed positive cases of Covid-19 infections available in the web site of the Italian “Ministero della Salute” ^1^, World Health Organization^2^ and Worldometer^3^, we found that they can be well approximated by a Cumulative Distribution Function (CDF) of the type of the Gauss Error Function, that is the integral of a normal, Gaussian distribution. Incidentally, such behavior is similar to the behavior of the incubation time of the seasonal influenza^4^. By positive cases we mean the positive cases actually diagnosed plus, for future days, the positive cases that we expect to be diagnosed. In fact it is well known among the virologists that the actual number of positive cases is much higher than the diagnosed ones^5^. However, it is assumed the diagnosed cases are a good statistical representation of the entire population of the positive cases. In Fig. 1, we report the result of our fit of the cumulative diagnosed positive cases in China. We have then applied such a CDF to study the evolution in time of the number of positive cases in Italy, in the attempt to possibly, statistically, predict the peak in the number of daily positive cases and the possible date of a substantial decrease in the number of daily positive cases. Finally, in section 4 we have performed a number of Monte Carlo simulations to get a more robust prediction of both the date of the peak in the number of positive cases each day and the date after which the number of new positive cases is below a certain threshold.

**Fig. 1.**
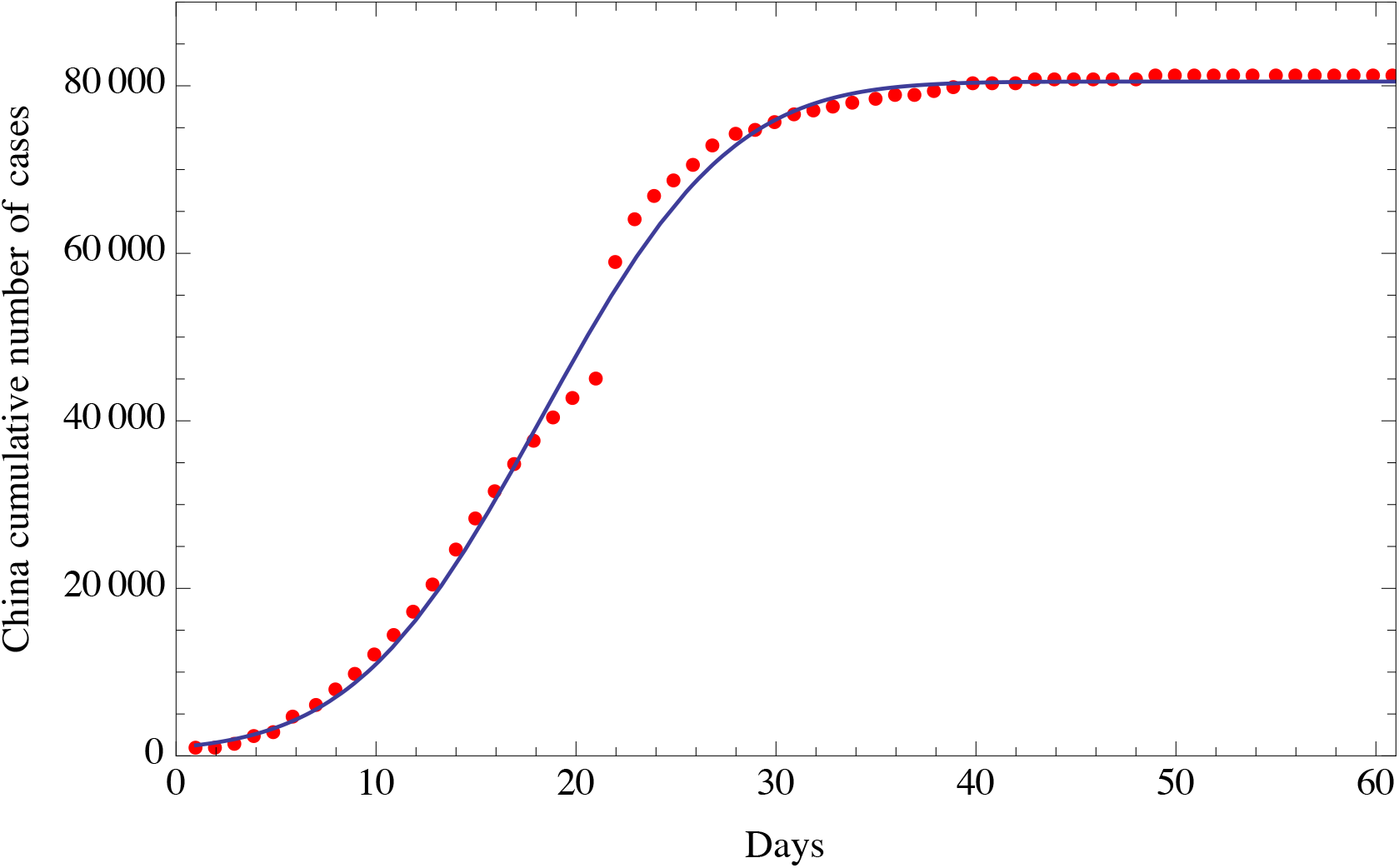
Fit of the cumulative number of diagnosed positive cases of Covid-19 in China (red dots) from January 22, 2020 to March 22, 2020 and the fitting function of the type of a Gauss Error Function solid line. The horizontal axis reports the days from the beginning of contamination; the vertical axis reports the cumulative number of diagnosed positive people.

## 2. Fit of cumulative diagnosed positive cases of Covid-19 in China

Based on the number of diagnosed positive cases of Covid-19 in China, we have fitted the cumulative numbers with a function of the type of the Gauss Error Function

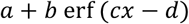

containing the four parameters *a, b, c, d*, that we have fitted using the available official data. Such a distribution is also observed in other studies of seasonal influenza^4^. The result of the fit is reported in Fig. 1, which shows the good level of the fit using such Gauss Error Function with those four parameters.

## 3. Fit and predictions of cumulative positive cases of Covid-19 in Italy

In this section we report the results of the fit of the cumulative diagnosed positive cases of Covid-19 in Italy using a function of the type of the Gauss Error Function, given in section 2. We obtained very similar results using a Logistic Function with four parameters (so for the sake of brevity we omit here to report that part). In Fig. 2 is shown the fit with data from February 15, 2020 to March 26, 2020. According to this fit, the flex, i.e. the point where the second derivative of the fit is becoming negative, that is the difference between two successive daily positive cases becomes negative, or in other words the point where there is a deceleration in the number of positive cases, is reached at March, 25, 2020. According to this fit, the date of a substantial reduction in the number of cumulative positive cases in Italy (below 100 cases) is April 22, 2020.

**Fig. 2.**
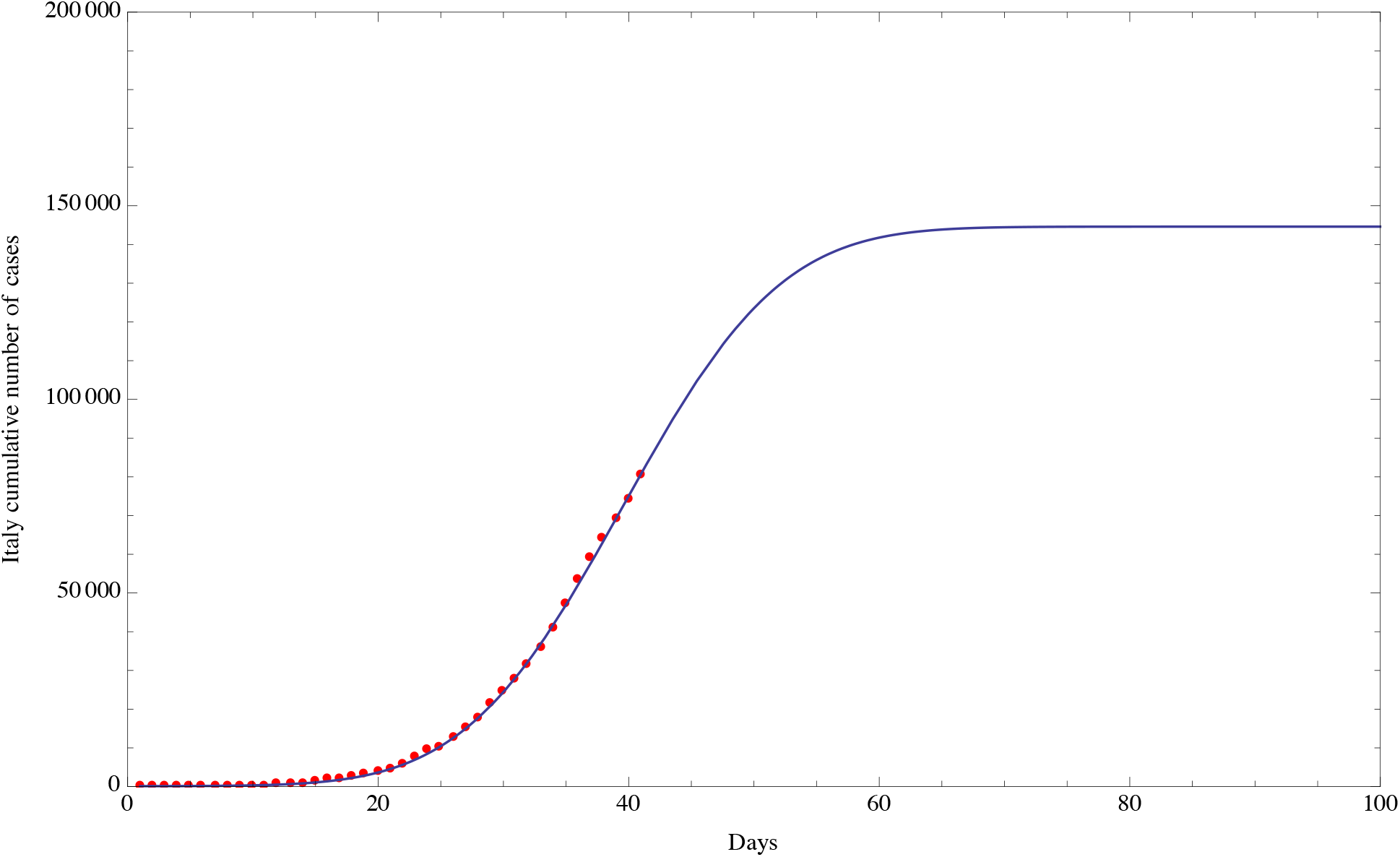
Fit of the Cumulative number of diagnosed positive cases of Covid-19 in Italy (red dots) from February 15, 2020 (included) to March 25, 2020 (included) using a fitting function of the type of the Gauss Error Function with four free parameters (solid line). A similar result was obtained with a fitting function of the type of the Logistic Function.

To evaluate the standard deviation relative to the date of the flex, we have used two methods. The first one is to fit the cumulative diagnosed positive cases in Italy using data from the beginning, February 15, 2020 (included) to March 16, 2020 (included), then from February 15, 2020 to March 17, 2020 and so on, until March 26, 2020 (included), thus getting 11 evaluations of the date of the flex. We then evaluated the standard deviation of these 11 points and we obtained a 1-sigma standard deviation of 3 days. The date of the flex has a quasi oscillating behavior whose amplitude is decreasing by increasing the final date of the analysis, i.e., increasing the number of points used in the fit. The 2-sigma (95.5% probability) and 3-sigma (99.7% probability) uncertainties in the day of the flex are then 6 days and 9 days respectively. In regard to the date of a substantial reduction in the number of cumulative positive cases in Italy (below 100 cases), the standard deviation is 6 days (68.2% probability), 12 days (95.5% probability) and 18 days (99.7% probability).

Using the second more robust method, we evaluated by a Monte Carlo analysis^6-8^ the day and the standard deviation of the flex and the day and the standard deviation of a substantial reduction in the number of diagnosed positive cases, as described in the next section 4.

## 4. Monte Carlo simulations of cumulative positive cases of Covid-19 in Italy

The Monte Carlo simulations^6-8^ have been designed to possibly take into account the measurement error in each daily number of the cumulative positive cases of Covid-19 in Italy. This error describes the uncertainty in the process of measuring the daily number of positive cases due to some fluctuations in the measurement procedures (such as a different number of performed daily tests of one day with respect to another day); of course this error does <u>not</u> describe the difference between the actual total positive cases and the diagnosed ones which can be very large^5^. However, the diagnosed cases are considered to be a representative sample of the population (i.e. of the total number of positive cases, which by the way is unknown). To get an estimate of the uncertainty in each daily number, we applied the following heuristic approach. We have assumed a measurement uncertainty in the total positive cases equal to 10% of each daily number (Gaussian distributed).

The second step was to generate a random matrix (*m*×*n*), where *n* (columns) is the number of days and *m* (rows) is a number of outcomes that we have chosen to be 150. Each number in the matrix is part of a Gaussian distribution with mean equal to 1 and sigma equal to 0.1 (i.e., 10% of 1), either row-wise and column-wise. In such a way each day is characterized by 150 simulated outcomes that allow to apply a statistical approach. The 150 outcomes represent a reasonably large number of simulated deviations from the official data according to the Central Limit theorem.

We then multiplied the nominal value of the diagnosed positive cases of the *j*-th day for the 150 numbers of the *j*-th column of the random matrix mentioned above. In such a way each day will be associated with a series of 150 numbers (Gaussian distributed with an uncertainty of 10% standard deviation) which simulate the statistical nature of the single datum. The index *j* will run from February 15, 2020 to March 26, 2020. Finally, from the daily data, we generated 150 series of cumulative diagnosed positive cases that allow to perform a statistical analysis.

In summary, starting from the *n* nominal values of the daily data, we generated *n* Gaussian distributions with 150 outcomes, with mean equal to the *n* nominal values and with standard deviation equal to 10%. Then, for each of the 150 simulations, the *n* residuals (corresponding to the cumulative positive cases of *n* days) were fitted with a four parameter function of the type of the Gauss Error Function and we then determined the date of the flex with such fitted function for each simulation. Using the fitted function we also determined the date at which the number of daily positive cases will be less than a certain threshold that we have, for example, chosen to be 100. Finally we calculated the standard deviation of these 150 simulations. In Fig. 3 and Fig. 4, we report the values (red dots) and the mean (horizontal line) of the Monte Carlo simulations respectively for the date of the flex and for the date of a substantial reduction in the number of daily positive cases.

**Fig. 3.**
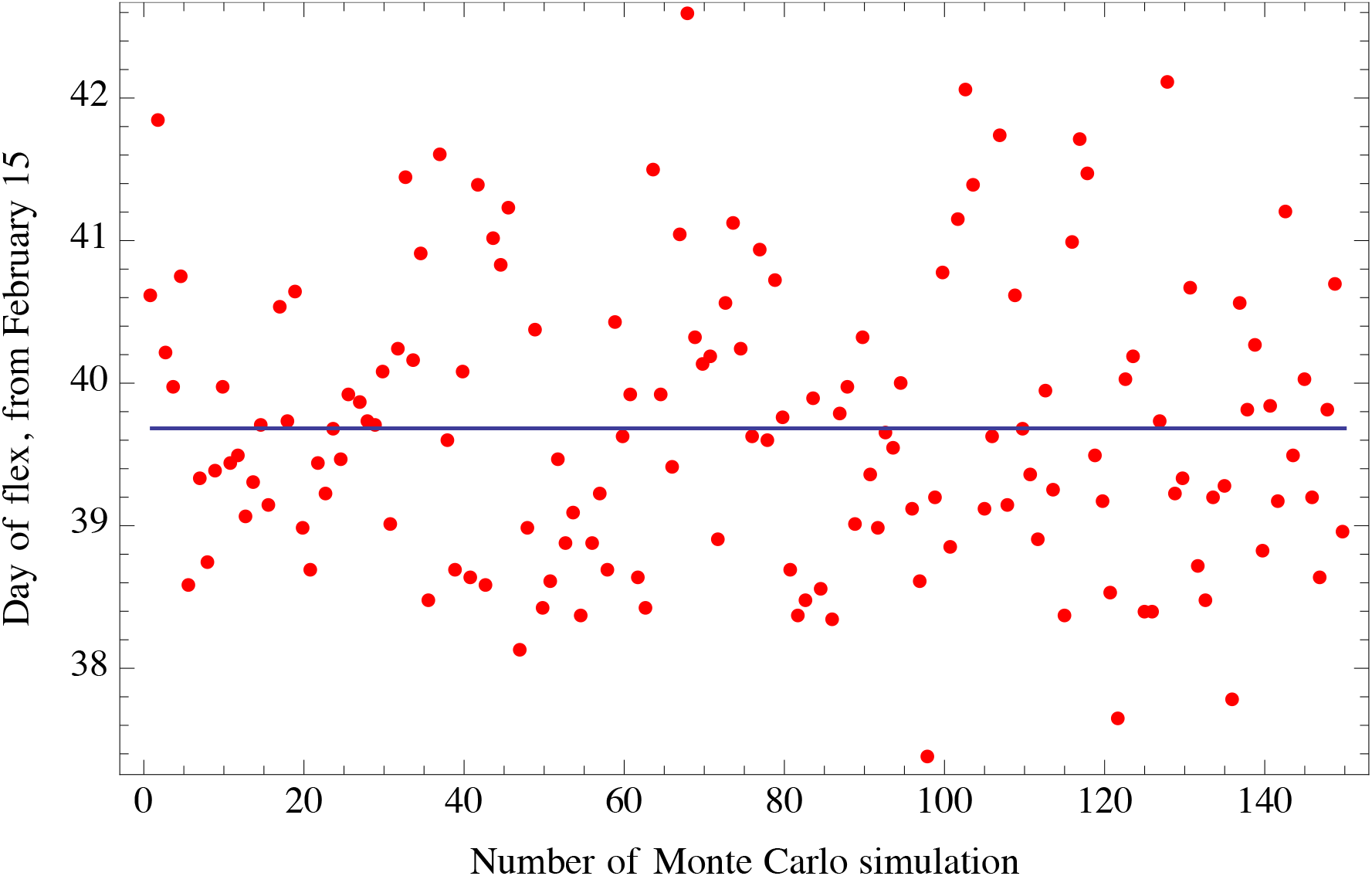
Monte Carlo simulations: each red dot corresponds to the day of occurrence of the flex (reduction in the number of daily cases, i.e., deceleration in the number of daily cases) obtained with each of the 150 Monte Carlo simulations. The vertical axis reports the number of days from February 15, 2020.

**Fig. 4.**
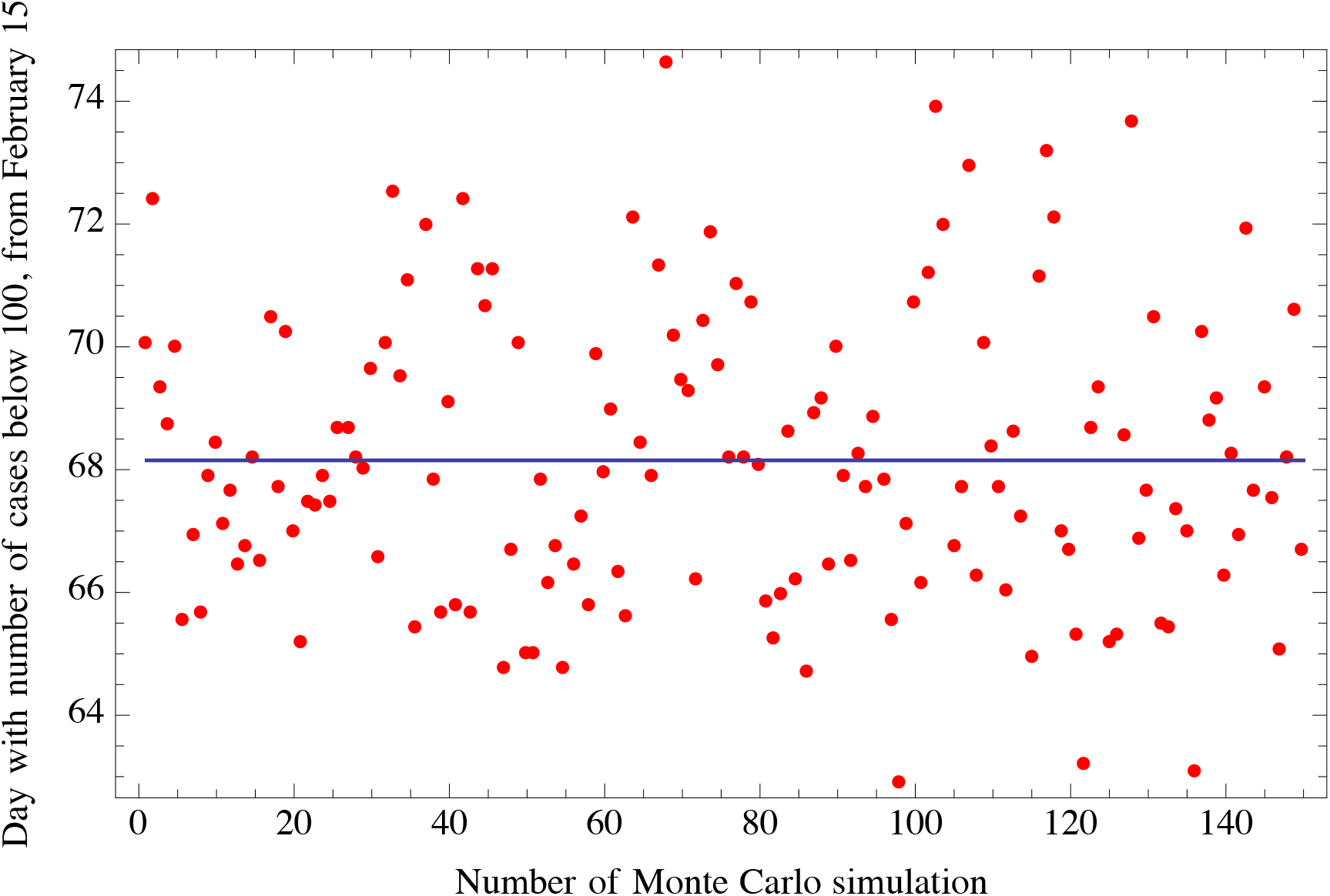
Monte Carlo simulations: each red dot corresponds to the day in which a substantial reduction in the number of daily cases (below 100) occurs. Each dot is obtained with each of the 150 Monte Carlo simulations. The vertical axis reports the number of days from February 15, 2020.

Using *n* = 41 days (i.e., the number of daily diagnosed positive cases up to March 26, 2020), we thus obtained a standard deviation (1-sigma) of 1 day for the date of the flex and of 2.3 days for the date in which a substantial reduction of the daily cases would be below 100. This result corresponds to a probability of 68.2% that the date of the flex will be at a certain date plus or minus 1 day and that the date of a substantial reduction of the number of cases will be at a certain date plus or minus 2.3 days. A 2-sigma standard deviation will give a more robust probability of 95.4% of the day of the flex and of the day of a substantial reduction in the number of cases. The 2-sigma values correspond to plus or minus 2 days for the day of the flex and plus or minus 4.6 days for the day of a substantial reduction.

## 5. Conclusions

Using the first 41 days of the number of cumulative diagnosed positive cases of Covid-19 in Italy (i.e., from February 15, 2020 to March 26, 2020) with a fitting function of the type of the Gauss Error Function with four free parameters (a distribution function which well fits the corresponding cumulative diagnosed positive cases in China), we obtained that the day of the flex (i.e., the day of deceleration in the number of daily positive cases) is in Italy, with a 95.4% probability, between March 23, 2020 and March 27, 2020; the 2-sigma uncertainty of +/- 2 days was obtained with a Monte Carlo simulation using the first 40 days. In regard to the day of a substantial reduction in the number of the daily positive cases (which, for example, we took to be less than 100), this day will be in Italy, with a 95.4% probability, between April 17, 2020 and April 27, 2020; the 2-sigma uncertainty of +/- 2 days was also obtained with the Monte Carlo simulation using the first 41 days. This predictions are statistical in nature and do not model the relevant factors of social distancing, and epidemiological and virology studies, which are outside the purpose of the present paper.

## Data Availability

The author are using data publicly available. The source of the data are listed in the references of the paper.

## Notes

### Competing Interest Statement

The authors have declared no competing interest.

### Funding Statement

The authors did not received any funding for the paper.

